# Hospital admissions and mortality for acute exacerbations of COPD during the COVID-19 pandemic: a nationwide study in France

**DOI:** 10.1101/2022.07.11.22277259

**Authors:** Jonas Poucineau, Tristan Delory, Nathanael Lapidus, Gilles Hejblum, Christos Chouaïd, Sophie Le Cœur, Myriam Khlat, the COVID-HOSP working group

**Affiliations:** French Institute for Demographic Studies (INED), Mortality, Health and Epidemiology Unit, Aubervilliers, 93322, France; Institute for Research and Information in Health Economics (IRDES), Paris, 75019, France; Annecy-Genevois Hospital Center, Annecy, France; Sorbonne Université, INSERM, Institut Pierre Louis d’Épidémiologie et de Santé Publique, Paris, France; Sorbonne Université, INSERM, Institut Pierre Louis d’Épidémiologie et de Santé Publique, AP-HP, Hôpital Saint-Antoine, Unité de Santé Publique, Paris, France; INSERM, IMRB (Clinical Epidemiology and Ageing Unit), Paris Est Créteil University, F-94010 Créteil, France; Pneumology Department, Intercommunal Hospital Center of Créteil, 40, Avenue de Verdun, F-94010 Créteil, France

## Abstract

**Background:** A global reduction in hospital admissions for acute exacerbations of chronic obstructive pulmonary disease (AECOPD) was observed during the first months of the COVID-19 pandemic. Large-scale studies covering the entire pandemic period are lacking. We investigated hospitalizations for AECOPD and the associated in-hospital mortality at the national level in France during the first two years of the pandemic.

**Methods:** We used the French National Hospital Database to analyse the time trends in (1) monthly incidences of hospitalizations for AECOPD, considering intensive care unit (ICU) admission and COVID-19 diagnoses, and (2) the related in-hospital mortality, from January 2016 to November 2021. Pandemic years were compared with the pre-pandemic years using Poisson regressions.

**Results:** The database included 565,890 hospitalizations for AECOPD during the study period. The median age at admission was 74 years (interquartile range 65–83), and 37% of the stays concerned women. We found: (1) a dramatic and sustainable decline in hospitalizations for AECOPD over the pandemic period (from 8,899 to 6,032 monthly admissions, relative risk (RR) 0.65, 95% confidence interval (CI) 0.65–0.66), and (2) a concomitant increase in in-hospital mortality for AECOPD stays (from 6.2% to 7.6% per month, RR 1.24, 95% CI 1.21-1.27). The proportion of stays yielding ICU admission was similar in the pre-pandemic and pandemic years, 21.5% and 21.3%, respectively. In-hospital mortality increased to a greater extent for stays without ICU admission (RR 1.39, 95% CI 1.35–1.43) than for those with ICU admission (RR 1.09, 95% CI 1.05–1.13). Since January 2020, only 1.5% of stays were associated with a diagnosis of COVID-19, and their mortality rate was nearly 3-times higher than those without COVID-19 (RR 2.66, 95% CI 2.41–2.93).

**Conclusion:** The decline in admissions for AECOPD during the pandemic could be attributed to a decrease in the incidence of exacerbations for COPD patients and/or to a possible shift from hospital to community care. The rise in in-hospital mortality is partially explained by COVID-19, and could be related to restricted access to ICUs for some patients and/or to greater proportions of severe cases among the patients hospitalized during the pandemic.

## 1. Introduction

Chronic obstructive pulmonary disease (COPD) is the third leading cause of death and affects over 390 million people worldwide.^1,2^ People living with COPD are prone to acute exacerbations of their symptoms, and severe exacerbations require hospital admission. If infected with COVID-19, they are at high risk of severity or mortality.^3,4^

A substantial decrease in the number of hospital admissions for acute exacerbation of COPD (AECOPD) during the first months of the COVID-19 pandemic has been reported in several countries,^5–17^ ranging from -42% to -48% among nationwide studies.^6–8,15,17^ This observation raises two critical concerns in terms of generalizability. First, whether this downturn pattern persisted over the entire pandemic period remains unknown, as most studies covered the first wave only,^6,7,12–16^ and no study spanning beyond February 2021 has yet been published. Second, to our knowledge, only five of the above-mentioned studies used national data,^6–8,15,17^ and these concerned relatively small countries (Wales, Scotland, Denmark, Slovenia and Norway), with the exception of South Korea, covering the first half of 2020 only.^15^ Furthermore, findings on in-hospital mortality trends for COPD patients during the pandemic are inconsistent. Two studies found an increased mortality in 2020 compared to pre-pandemic years, of respectively +53% and +130% but both were based on small numbers,^8,16^ while others did not find a significant difference.^11,12,14^ To date, time trends in in-hospital mortality have not been analysed with large-scale and exhaustive data. Finally, while most studies found a stable proportion of AECOPD stays involving intensive care units (ICU) admission in 2020 compared to previous years,^11,14,16^ a cohort study in Denmark found a 36% decrease in the risk of admission to an ICU in COPD patients.^7^

France has been heavily affected by the COVID-19 pandemic, with over 31 million confirmed cases and 146,000 deaths as of July 2022.^18^ Numerous health measures have been implemented, including stringent lockdowns and travel restrictions, likely to affect care for non-COVID-19 disorders. During lockdowns, hospitals suspended most ‘non-essential’ activities to focus on managing COVID-19 cases.

Based on exhaustive hospitalization data at the national level, the present study describes how hospitalizations for acute exacerbations of COPD (AECOPD) and the associated in-hospital mortality varied in France throughout the first two years of the pandemic. The specific objectives of the study were to (1) study trends in admissions and mortality by demographic characteristics, (2) compare the observed trends with those estimated in the absence of a pandemic, (3) explore possible associations between COVID-19 diagnosis, ICU admission and in-hospital mortality, (4) suggest interpretations of the results based on the literature and the context in France during the pandemic.

## 2. Materials and methods

This observational study was conducted according to STROBE guidelines.^19^ The study is based on the French National Hospital Database (*Programme de Médicalisation des Systèmes d’Information*; PMSI) from the National Health Data System (*Système National des Données de Santé*; SNDS), an exhaustive and nationwide administrative health claims database, covering 99% of the French population, i.e. over 66 million people, from birth or immigration until death or emigration. The PMSI covers all private and public medical, obstetric and surgery hospitalizations.^20^ Patients can be tracked individually with a unique and anonymous identifier. All hospital stays are assigned a primary diagnosis (PD) and possibly one or more associated diagnoses (AD), coded according to the International Classification of Diseases, 10th revision (ICD-10).We identified hospital stays for AECOPD using an algorithm based on ICD-10 codes, validated by the French Institute for Public Health, and adapted by Cavailles et al.^21^ This algorithm is based on a narrow definition of AECOPD to avoid overlap with asthma or other chronic pulmonary condition. We included patients aged 40 years or older with compatible ICD-10 codes:

- J44.0 or J44.1 as PD,
- or J96.0 as PD and (J43 or J44) as AD,
- or (J09-18 or J20-22) as PD and (J43 or J44) as AD,
- or (J43 or J44) as PD and (J44.0 or J44.1 or J96.0 or J09-18 or J20-22) as AD.

This algorithm excludes stays lasting fewer than two days, unless patients died during hospitalization, in order to eliminate scheduled hospitalizations for check-up examinations.

Among the study population, we identified stays with an associated diagnosis of COVID-19, using the ICD-10 code U07.1. In the PMSI, the code U07.2 has not been created and all COVID-19 diagnoses were grouped within the code U07.1.

The study period extended from January 2016 to November 2021 and was divided into two main periods: one further referred to as the “pre-pandemic years” including data from 2016 to 2019 as the reference, and the other as the “pandemic years” including data from January 2020 to November 2021. During the pandemic years, three sub-periods, representing broadly the first three waves of the pandemic in France, were especially investigated: March to June 2020, September to December 2020, and January to May 2021.

The main outcomes of this study were the time trends in (1) monthly incidences of hospitalizations for AECOPD and (2) the corresponding in-hospital fatality rate. Admissions to intensive care units (ICU), COVID-19 diagnoses and their relation to in-hospital mortality were also investigated. Analyses were stratified by sex and age groups (40–49 years, 50–59, 60–69, 70–79, and 80 or older). Relative risks (RR) were estimated by comparing each sub-period of the pandemic to the same months in 2016–2019, for both outcomes. Poisson regressions of the monthly counts of admissions and in-hospital deaths were performed, adjusting on sex, with random intercepts for month and age group, and using national census data and AECOPD admissions as offsets. Expected numbers of admissions and in-hospital deaths in 2020-2021 in the absence of a pandemic were obtained by Poisson regressions using a similar method, adding a sinusoidal component to account for seasonality. To estimate the expected numbers of in-hospital deaths in 2020-2021, 2016-2019 in-hospital mortality rates were applied to observed stays in 2020-2021. Deaths attributable to COVID-19 were estimated as the difference between observed and expected deaths in 2020-2021

Sensitivity analyses were conducted to test the variability of our results according to the inclusion or exclusion of stays lasting fewer than two days, and the definition of AECOPD admission (broad vs. narrow).The broad definition of AECOPD was based on an algorithm developed by Fuhrman et al.,^22^ which includes all stays with an ICD-10 code J44 or J43 as primary diagnosis and those with the same codes or combination of codes as described above, regardless of the type of diagnosis (primary or associated).

No missing data had to be processed, as all variables of interest were correctly filled in. The study was conducted in accordance with relevant international and French regulatory requirements, including the dedicated permanent authorization to use PMSI granted to the French Institute for Demographic Studies (INED) by Decree No. 2016-1871 of December 26, 2016 on the processing of personal data. The study was registered on the European Network of Centres for Pharmacoepidemiology and Pharmacovigilance (ENCePP) registry under the number “EUPAS40492”.

The study was funded by the grant PREPS-20-0163 (*Programme de recherche sur la performance du système des soins*, French Ministry of Solidarity and Health).

## 3. Results

### 3.1. Hospital admissions

From January 1, 2016 to November 30, 2021, 321,013 individuals were hospitalized for AECOPD, with a total of 565,890 corresponding admissions (Table 1). The median age at admission was 74 years (interquartile range 65–83), and 209,290 (37%) concerned women. There was a seasonal pattern in monthly incidence of hospitalizations for AECOPD during the reference pre-pandemic years, with the highest levels observed from December to March and the lowest levels observed from July to September. Such a pattern did not occur during the pandemic years (Figure 1).

**Table 1.** Mortality and ICU admission rates for AECOPD stays (2016-2021) AECOPD, acute exacerbation of chronic obstructive pulmonary disease; ICU, intensive care unit; AD, associated diagnosis.

|  | Stays | Deaths | Mortality rate | ICU admission rate |
| --- | --- | --- | --- | --- |
| **Years 2016-2019** | 427165 | 26562 | 6.2% | 21.5% |
| **Years 2020-2021\*** | 138725 | 10322 | 7.4% | 21.3% |
| with Covid-19 AD | 2105 | 424 | 20.1% | 32.4% |
| without Covid-19 AD | 136620 | 9898 | 7.2% | 21.1% |

\* December 2021 data are not included

**Figure 1.**
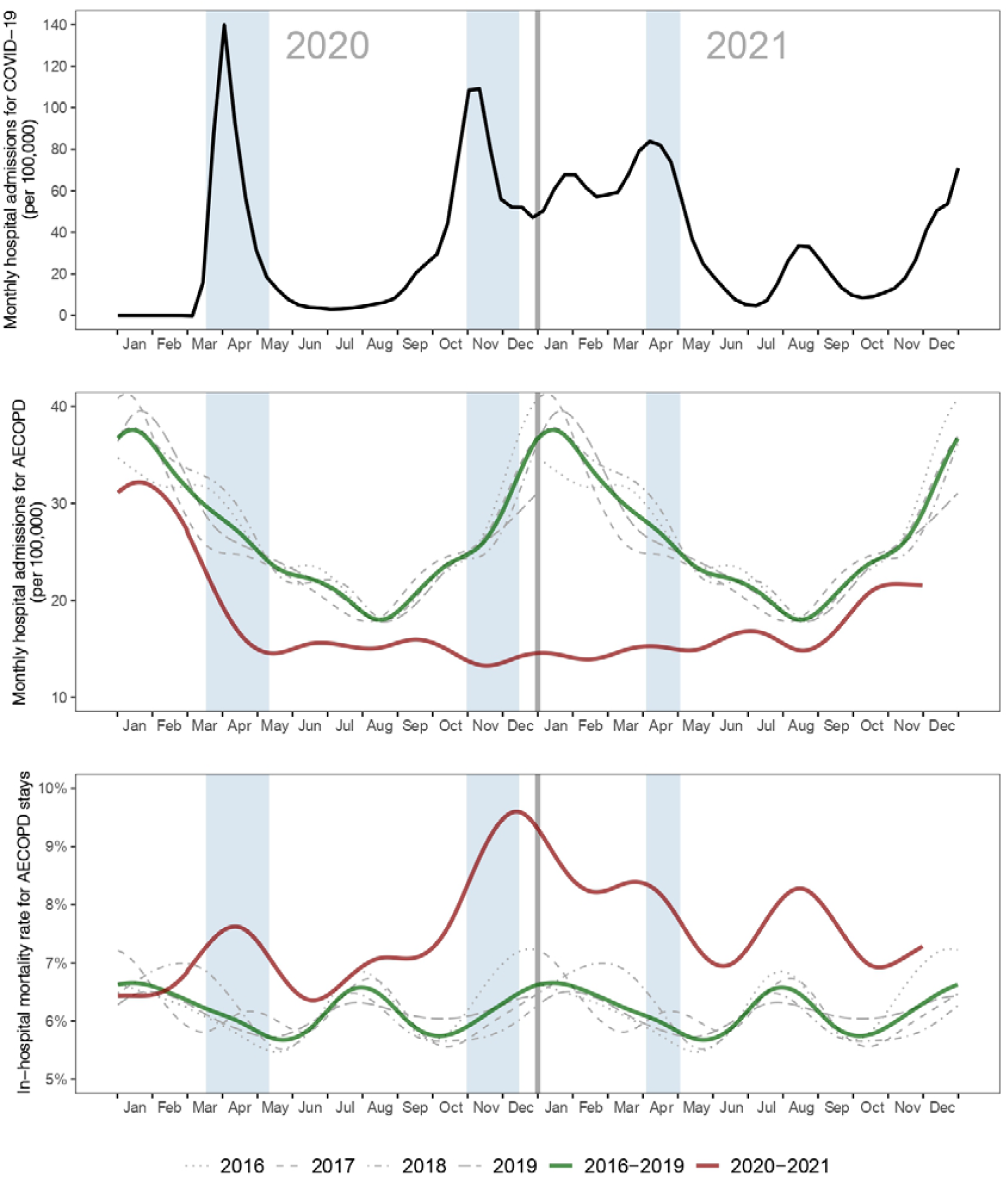
Time trends from January 2020 to November 2021 for **(A)** monthly incidence of COVID-19 hospital admissions, **(B)** monthly incidence of AECOPD hospital admissions and **(C)** in-hospital fatality rates for AECOPD. All curves were smoothed (cubic splines). In panels B and C, values for years 2016 to 2019 were used as references and the 2016–2019 curve averages them. Reference values have been duplicated on both sides of the graph to compare them with those of 2020 to 2021.Shaded areas correspond to the three lockdown periods in France. AECOPD, acute exacerbation of chronic obstructive pulmonary disease.

Overall, the average monthly incidence of admissions for AECOPD decreased by 35% during the pandemic years compared to pre-pandemic years (from 8,899 to 6,032 admissions per month, RR 0.65, 95% CI 0.65–0.66; Table 2). The gap widened across the three pandemic sub-periods, from a 34% decrease in the first sub-period (RR 0.66, 95% CI 0.65–0.67), to 45% in the second (RR 0.55, 95% CI 0.55–0.56), and 53% in the third (RR 0.47, 95% CI 0.46–0.47). Thereafter, the incidence of hospitalizations for AECOPD remained low until October 2021, when it started to rise slightly again. This downward trend was observed across all age groups, and was enhanced in the group of patients aged 80 or older versus others (RR 0.84, 95% CI 0.83–0.85). Trends were similar for in men’s and women’s during the pandemic years.

**Table 2.**
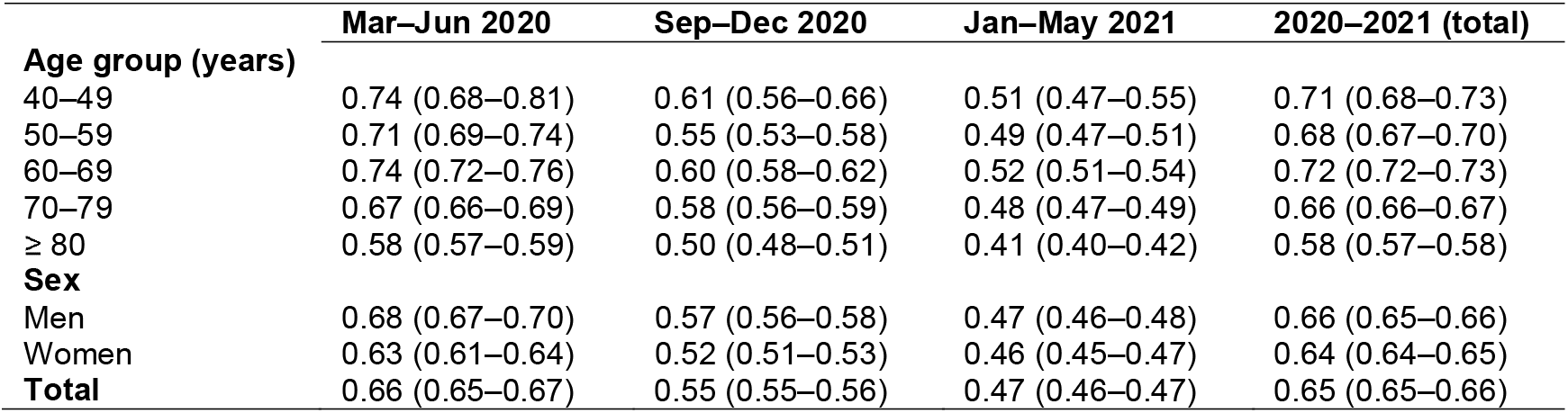
Relative risk of hospital admission for AECOPD for different pandemic periods, compared with the same months over the reference years, by age group and sex. Relative risks (95% confidence intervals). AECOPD, acute exacerbation of chronic obstructive pulmonary disease.

From January 2020 to November 2021, 2,105 hospitalizations for AECOPD also had a COVID-19 associated diagnosis, accounting for 1.5% of total admissions (Table 1). The proportion of AECOPD stays involving an admission to an ICU was similar in the pre-pandemic and pandemic years (21.5% and 21.3%, respectively). Exacerbations associated with COVID-19 more frequently resulted in ICU admission than those without (32.4% vs. 21.1%, RR 1.58, 95% CI 1.46–1.70).

Whereas 158,725 hospital stays for AECOPD were observed during the pandemic years, we estimated that in the absence of a pandemic, a total of 58,691 additional stays would have been expected in the same period (+37%): 27,095 (95% CI 22,093–32,296) stays in addition to the 76,000 stays observed in 2020, and 31,596 (95% CI 25,334–38,325) stays in addition to the 62,725 stays observed in 2021.

### 3.2. In-hospital mortality

The average monthly in-hospital mortality rate for AECOPD stays increased by 24% during the pandemic years (from 6.2% to 7.6% fatal issues, RR 1.24, 95% CI 1.21–1.27), reaching a peak at 9.8% in December 2020 (see Figure 1 and Table 3). The increase in in-hospital mortality was observed across all age groups during the pandemic years. This trend was attenuated in the group of patients aged 80 or older compared with others (RR 0.92, 95% CI 0.88–0.97). Trends in men’s and women’s in-hospital mortality were similar during the pandemic years, except for the second sub-period, when men had higher excess in-hospital mortality.

**Table 3.** Relative risk (95% confidence interval) of in-hospital death for AECOPD stays for different pandemic periods, compared with the same months over the reference years, by age group and sex. Relative risks (95% confidence intervals). AECOPD, acute exacerbation of chronic obstructive pulmonary disease.

|  | Mar–Jun 2020 | Sep–Dec 2020 | Jan–May 2021 | 2020–2021 (total) |
| --- | --- | --- | --- | --- |
| **Age group (years)** |  |  |  |  |
| 40–49 | 2.41 (1.39–4.20) | 1.98 (1.06–3.68) | 1.42 (0.79–2.56) | 1.48 (1.11–1.97) |
| 50–59 | 1.03 (0.79–1.35) | 1.29 (0.98–1.69) | 1.20 (0.94–1.53) | 1.15 (1.03–1.29) |
| 60–69 | 1.22 (1.08–1.37) | 1.43 (1.26–1.62) | 1.42 (1.27–1.59) | 1.28 (1.21–1.35) |
| 70–79 | 1.31 (1.19–1.44) | 1.55 (1.41–1.71) | 1.45 (1.33–1.58) | 1.31 (1.25–1.37) |
| ≥ 80 | 1.17 (1.08–1.26) | 1.34 (1.24–1.45) | 1.29 (1.21–1.38) | 1.20 (1.16–1.24) |
| **Sex** |  |  |  |  |
| Men | 1.23 (1.15–1.31) | 1.47 (1.38–1.56) | 1.37 (1.29–1.45) | 1.25 (1.22–1.29) |
| Women | 1.20 (1.09–1.32) | 1.31 (1.19–1.45) | 1.33 (1.23–1.44) | 1.23 (1.18–1.28) |
| **Total** | 1.22 (1.15–1.28) | 1.42 (1.34–1.49) | 1.35 (1.29–1.42) | 1.24 (1.21–1.27) |

Stays associated with a COVID-19 diagnosis had a substantially higher mortality rate than those without (20.1% vs. 7.2%, RR 2.66, 95% CI 2.41–2.93). Yet, stays without COVID-19 still had a significant increase in in-hospital mortality during the pandemic years compared to pre-pandemic years (from 6.2% to 7.2%, RR 1.21 95% CI 1.18–1.24).

In-hospital mortality rates for stays involving an ICU admission increased slightly during the pandemic years in comparison with pre-pandemic years (from 12.9% to 13.8%, RR 1.09, 95% CI 1.05–1.13). In-hospital mortality rates for stays not involving an ICU admission increased to an even greater extent (from 4.4% to 5.8%, RR 1.39, 95% CI 1.35–1.43).

We estimated that, among the 10,322 deaths observed for AECOPD stays during the pandemic years, 1,974 deaths (19%) were in excess: in the absence of a pandemic, 862 (95% CI 681–1,026) fewer deaths would have been expected in 2020 and 1,112 (95% CI 940–1,272) in 2021 compared to the 5,487 and 4,835 deaths observed, respectively. Among stays associated with COVID-19, we estimated that 291 deaths occurred in excess during 2020-2021, including 151 (95% CI 148–154) in 2020 and 140 (95% CI 137–143) in 2021. Overall, since the pandemic started, COVID-19 accounted for 14.8% (95% CI 12.9%–17.6%) of the estimated excess deaths for AECOPD stays.

### 3.3. Sensitivity analysis

There was a greater proportion of stays shorter than two days among AECOPD stays during the pandemic years compared to pre-pandemic years (14% and 10%, respectively). Nevertheless, similar trends in AECOPD admissions and in-hospital mortality were obtained when stays shorter than two days were included in the analyses. Likewise, results based on the broad definition of AECOPD were very similar to those reported above (data not shown).

## 4. Discussion

The present study is, to our knowledge, the largest study to investigate in detail AECOPD hospitalizations during the 2020 and 2021 pandemic years, compared with the 2016-2019 pre-pandemic years. To date, the largest study in the literature contained fewer than 17,000 hospitalisations.^7^ Based on the corresponding 565,890 hospital stays for AECOPD, the major findings of this study are (1) a sharp decline in the number of AECOPD hospitalizations during the 2020-2021 period, and concomitantly, (2) a substantial increase in in-hospital mortality.

A reduction in hospitalizations for AECOPD during the first months of the pandemic has been documented in the literature, with estimates ranging from -42% to -48% among nationwide studies.^6–8,15,17^ In this study, we show that this phenomenon persisted far beyond the first pandemic wave, with a global decline of -35% hospital admissions when considering the whole 2020-2021 pandemic years, and nearly 60,000 fewer stays than expected in the absence of a pandemic. In contrast, hospitalizations for causes other than COVID-19 reduced by only 13% in France in 2020 compared to 2019,^23^ indicating that the decline was steeper for AECOPD-related hospitalizations.

Several factors may have contributed to this sharp decline. First, public health measures are likely to have had protective effets on the onset of COPD exacerbations, mainly through a substantial reduction in the circulation of viral respiratory infections (VRI), the most common triggers of AECOPD.^24–26^ Evidence supports this interpretation. A study in Singapour showed a reduction in VRI-associated AECOPD during the first months of the pandemic,^13^ while in Hong Kong, an increase in the use of face masks was associated with a decrease in AECOPD.^12^ Another study in the United States showed a positive correlation between COPD admissions and community viral burden.^11^ The present study shows that in France, in contrast with what was observed during the pre-pandemic years, the number of hospital admissions for AECOPD did not increase during the winter months of the pandemic, a season when the circulation of VRI is usually high. Furthermore, a considerable increase in flu vaccination coverage was observed in France in 2020 compared with 2019.^27^ This suggests a real reduction in AECOPD due to decreased exposure to VRI, leading to a reduction in this population’s need for hospital care.^7^ Moreover, COPD patients have demonstrated strict compliance with health regulations during the pandemic in some countries, along with greater adherence to their treatments.^10,28,29^ Finally, the reduction of air pollution observed in France in 2020 likely constituted another protective effect on the onset of AECOPD.^30,31^

The pandemic context, however, also had negative effects on COPD patients’ health, with a deterioration in their mental health due to isolation^14,29,32,33^ and a reduction in their physical activity.^14,29^ These aspects represent risk factors for AECOPD and might therefore dampen slightly the above-mentionned protective effets on COPD patients.

On the other hand, changes in health-seeking patterns in COPD patients during the pandemic might have contributed to the decrease in hospital admissions for AECOPD. Several studies have reported hospital avoidance behaviours for fear of becoming infected with COVID-19,^14,33,34^ and access to hospital care was reduced by overcrowding during the pandemic. These aspects may have led to a shift from hospital care to ambulatory care. Some patients who would have normally been admitted to hospitals might have instead been treated by ambulatory care. Furthermore, the development of remote care (telemedicine) and of self-medication might also have contributed to a lesser use of hospital care for COPD patients.^29,33^ This hypothesis is not supported by a study from Wales in which a reduction in primary care consultations for AECOPD was observed in 2020 compared with previous years.^6^ In contrast, another study from England reports an increase in community managed exacerbations in 2020 compared with 2019, but this study is limited to a single secondary care clinic.^29^

Finally, another hypothesis to explain the reduction in AECOPD-related hospitalizations could be an excess mortality among the patients with prevalent severe COPD. Some of these patients may have died because of COVID-19 or reduced access to health-care. However, this assumption is not supported by nationwide studies from Denmark and Slovenia, where all-cause mortality was reported to decrease in COPD patients, regardless of severity.^7,8^

Our second major finding is a 24% increase in the proportion of in-hospital deaths for AECOPD stays during the pandemic, with nearly 2,000 estimated excess deaths. There could be several reasons for this observation. First, stays with a COVID-19 diagnosis had significantly higher in-hospital mortality and ICU admission rates compared to those without COVID-19. These findings are in line with several studies in the literature showing higher severity and mortality for COPD patients when infected with COVID-19.^3,4^ Given that the proportion of stays with a COVID-19 diagnosis was low (1.5%), the contribution of COVID-19 to the excess mortality observed for AECOPD stays was limited (15%). Second, a number of stays that would have required ICU admission may not have benefited from it due to overcrowding during the critical phases of the pandemic, thereby contributing to the excess in-hospital mortality. Indeed, the increase in in-hospital mortality was significantly lower for stays with ICU admission compared to stays without ICU admission. Yet, the proportion of stays involving an admission to ICU remained stable from 2016 to 2021 (∼21%). Third, there might have been a shift of the clinical profile of the admitted cases, with a higher proportion of severe cases in 2020 and 2021 compared with previous years. This could be due to some mild or moderate cases resorting to ambulatory care to avoid COVID-19 infection in hospital settings, and/or to delays in healthcare for some patients during the first wave, leading to increased severity. In other contexts, though, the decrease in admissions during the pandemic was greater for severe AECOPD compared with mild or moderate disease.^8,10^

Finally, the case of patients aged 80 years or older seems paradoxical. While they exhibited a greater decrease in hospitalizations for AECOPD during the pandemic period compared with other age groups, they did not experience greater excess in-hospital mortality. One explanation could be excess out-of-hospital mortality for this subpopulation, with some severe cases dying without seeking hospital care.

This study has several limitations. The interpretation of the findings is limited, as we lacked information in order to establish causal links between the observed trends and the suggested explanatory factors. Further explorations of the issues at stake would necessitate granulated information on clinical presentation at admission, prevention measures and prevailing air pollution. As regards to hospital deaths attributable to COVID-19, although the database is very robust, there could have been an underestimation of COVID-19 cases due to the lack of access to PCR testing at the beginning of the pandemic. Besides, the causes of death were not available and we are therefore unable to know the exact proportion of deaths related to COVID-19.

In conclusion, our findings provide reliable long-term trends in AECOPD hospital admissions and in-hospital mortality at a national level, reflecting sustained direct and indirect effects of the pandemic. Further analyses on individual-level data are needed to explore the trajectories of COPD patients at risk of hospitalization. Several crucial questions remain open regarding patterns of AECOPD, out-of-hospital mortality in COPD patients, and possible shifts in out-of-hospital health-care use.

## Data Availability

The analysis described in this manuscript was performed in a national database (Programme de Medicalisation des Systemes d Information; PMSI) belonging to a third-party, namely the French Ministry of Health. Access to this database is available to researchers on reasonable demand, after proper training and upon request to the relevant agency, the Caisse Nationale d Assurance Maladie (CNAM), on condition that the analyses envisaged have been authorised by the French Data Protection Agency (Commission nationale de l Informatique et des Libertes, CNIL).

The **COVID-HOSP working group** includes: Tristan Delory, Centre Hospitalier Annecy-Genevois, Annecy, France; Fanny Duchaine, IRDES, Paris, France; Maude Espagnac, IRDES, Paris, France; Coralie Gandré, IRDES, Paris, France; Gilles Hejblum, INSERM, Paris, France; Myriam Khlat, INED, Aubervilliers, France; Nathanael Lapidus, INSERM, Paris, France; Sophie Le Cœur, INED, Aubervilliers, France; Elhadji Leye, INSERM, Paris, Jonas Poucineau, INED, Aubervilliers, France.

## Contributors

The study was initiated by NL, TD and GH. Data extraction and management were performed by JP. JP, NL, TD and GH contributed to data analysis and conception of figure and table. JP prepared the first draft. JP, NL, TD, GH, CC, SL and MK reviewed the manuscript. All authors contributed to the concept and design of the study, critical revision of the manuscript, and approved the final version. Members of the working group participated in interpreting and discussing the results.

## Data sharing

The analysis described in this manuscript was performed in a national database (*Programme de Médicalisation des Systèmes d’Information;* PMSI) belonging to a third-party, namely the French Ministry of Health. Access to this database is available to researchers on reasonable demand, after proper training and upon request to the relevant agency, the *Caisse Nationale d’Assurance Maladie* (CNAM), on condition that the analyses envisaged have been authorised by the French Data Protection Agency (*Commission nationale de l’Informatique et des Libertés*, CNIL).

## Acknowledgements

We thank Christopher Leichtnam for his editorial work. The study was funded by a grant from the French Ministry of Solidarity and Health (PREPS-20-0163). The funder had no role in study design, collection, analysis, and interpretation of the data, writing of the study report, decision to submit the report for publication.

